# Reporting trends and seasonality in the World Health Organization’s Disease Outbreak News (DONs) across 1996-2023

**DOI:** 10.1101/2025.06.17.25329744

**Authors:** Kellerine Quah, Jingtong Jin, Charlotte Christiane Hammer

## Abstract

The Disease Outbreak News (DONs), which are published by the World Health Organization, are used in outbreak surveillance, management and research as official reports of disease outbreaks worldwide. This study analyzed datasets of the DON created by Carlson et al. (2023) and Weets et al. (2025) to understand reporting behavior in DONs published across 1996-2023. Time series analysis was conducted by disease transmission group and geography. Interrupted time series analyses were conducted to investigate changes in reporting behavior before and after the 2003 SARS-CoV-1 outbreak, and before and during the COVID-19 pandemic. We observed an annual, 12-month seasonality in reporting across all DONs. By disease transmission group, Airborne and Droplet transmission-related DONs and Vector-borne transmission-related DONs exhibited 12-month seasonality in reporting. Faecal-oral and FWB transmission-related DONS exhibited 9-month seasonality in reporting. Findings from our interrupted time series analyses revealed that during the COVID-19 pandemic, reporting across all DONs decreased, with a step-change difference of −3.73 reports at baseline comparing negative binomial regression models before and during the pandemic. In addition, after the 2003 SARS-CoV-1 outbreak, reporting of Airborne and Droplet transmission-related DONs increased, with a step-change difference of 3.77 reports at baseline comparing negative binomial regression models before and after the outbreak. As a collection of press releases which are selectively published by the WHO, the DON is not a complete representation of global disease outbreaks, and researchers should be aware of these reporting patterns and biases when using the DONs in outbreak surveillance and global health research.

## Introduction

The Disease Outbreak News (DONs) are news reports of infectious disease outbreaks globally, which have been publicly disseminated online by the World Health Organization since 1996 [1]. With an estimated 2.6 million visits per year, the DONs web page is frequently visited, as one of the main platforms used by the WHO to communicate verified information on disease outbreaks to the public [2]. The DONs are highly relevant for disease surveillance, and can influence outbreak prevention, control and management [3, 4]. In global health research, previous uses of the DONs span epidemiology, economics and policy, including analyses of the timeliness of outbreak reporting [5–7], the analysis of economic indicators through measuring outbreak events [7], and qualitative analyses to understand the prioritization of infectious diseases by policymakers [8, 9].

The WHO does not publish every disease outbreak in the DON, but selects specific outbreaks based on the following criteria: (i) the ‘need for assistance from the international community’, (ii) ‘potential public interest’, (iii) whether ‘information on the event has already been reported publicly’, and (iv) the ‘need to disseminate authoritative and independent information’ [2]. As a result of this selection process, overreporting and underreporting of disease outbreaks may have occurred, which would have implications on global disease surveillance and outbreak management. For example, the underreporting of disease outbreaks may have decreased public awareness, minimized public health action and reduced potential funding for diseases and countries in need of international attention and assistance [10–12]. By analyzing reporting behavior in the DON, we can elucidate potential biases and highlight areas for improvement, to benefit the overall impartiality and quality of reporting in the DON.

## Methods

### Dataset

We merged a structured dataset of DON reports between 1996 and 2019 developed by Carlson et al. (2023) [13], and a dataset of DON reports between 2020 and 2023 by Weets et al. (2025) [14], to form a dataset of DON reports published from 22 January 1996 to 31 December 2023.

Both datasets are publicly accessible and available online on GitHub, at https://github.com/cghss/dons and https://github.com/cghss/dons2/tree/main. Individual DON reports may be accessed directly from the WHO website, at https://www.who.int/emergencies/disease-outbreak-news. Data cleaning was conducted and information on the process of removing duplicates is provided in the Supplementary Material. For data synthesis and analysis, R (version 4.2.2) using RStudio (version 2022.07.2+576) was used. The list of R packages used has been included in S1 Table.

To comprehensively analyze reporting patterns in the DON, we selected variables providing general information about the DON, such as “DONid”, “Headline”, “ReportDate”, “Country” and “DiseaseLevel1”. The definitions of selected variables are provided in S5 Table.

### Disease Classification

By adapting a classification of communicable diseases by Webber (2010) [15], we classified 78 diseases featured in the DON into 7 disease transmission groups, as defined in the Table. Further information on the process of disease classification and the full list of diseases are included in the Supplementary Material.

**Table.** Definitions of disease transmission groups for classifying diseases in the Disease Outbreak News (DONs).

| Disease Transmission Group | Definition |
| --- | --- |
| Airborne and Droplet | Diseases transmitted through small and/or large respiratory droplets |
| Faecal-oral and FWB | Diseases transmitted through the faecal-oral route, and food-borne and water-borne diseases |
| Fluid-borne | Disease transmitted through infectious bodily fluids, including blood, semen, vaginal secretions, cerebrospinal fluid, etc. |
| STDs and Skin Contact | Includes sexually transmitted diseases and diseases of skin contact |
| Vector-borne | Diseases transmitted by vectors, including mosquitoes, sandflies, fleas, ticks, etc. |
| Zoonoses | Diseases transmitted from non-human animals to humans |
| Other | Includes diseases which have not been specified and/or diseases for which the transmission route is not well-defined |

### Hemisphere Split

As the Northern and Southern hemispheres experience opposite seasons at the same time of the year, categorizing the reports by hemisphere can help capture the differences in disease dynamics and is hence useful for analyzing seasonality in the DONs. We classified 173 countries, as well as wider geographical regions reported in the DON, to the Northern or Southern hemisphere by geographical location and analyzed reporting patterns for the Northern and Southern hemispheres.

For cases with a clear outbreak location, we used the Google Maps API via the ‘ggmap’ package in R to obtain latitude coordinates. Latitude coordinates > 0 were classified as the Northern hemisphere, and latitude coordinates < 0 were classified as the Southern hemisphere. Report locations which were not recognized by the Google Maps API were classified manually. For example, the “African Meningitis Belt” was classified under the Southern Hemisphere.

### Time Series Analysis

Time series analysis was used to analyze patterns and trends in the DONs across time [16]. All statistical tests were conducted at a significance level of 5%. Linear regression, Poisson regression and negative binomial regression were used to model the data. To model seasonality, we added sine and cosine terms to the model, for which 3 parameters were required: (i) amplitude, (ii) period, and (iii) time. A periodogram was plotted to calculate the significance of different frequencies. Residual analysis was performed to check for overdispersion. Final model selection was determined using Akaike Information Criterion (AIC) and a likelihood ratio test [17].

### Interrupted Time Series Analysis

We conducted two interrupted time series analyses to understand the impacts of significant events on reporting patterns in the DON [18]. First, we analysed the impact of the COVID-19 pandemic on reporting across all DONs, comparing models before and during the COVID-19 pandemic. The start point of the COVID-19 pandemic was defined as 5 January 2020, the date of the first DON report on COVID-19 [19]. The end point of the COVID-19 pandemic was defined as 5 May 2023, when WHO released a statement declaring that COVID-19 no longer constituted a PHEIC [20]. Next, we compared models of reporting of Airborne and Droplet transmission-related DONs, before and after the 2003 SARS-CoV-1 outbreak. The start point of the SARS-CoV-1 outbreak was defined as the first WHO announcement of the outbreak (11 February 2003) [21], and the end point of SARS-CoV-1 outbreak was defined as the date on which the WHO declared the SARS-CoV-1 outbreak to be contained (5 July 2003) [22, 23]. The step-change difference of both models was calculated to obtain the difference in report count at baseline. Further information on the calculation of step-change difference is included in the Supplementary Material.

## Results

### Descriptive Temporal Analysis by Transmission Group and Hemisphere

Overall, there are a total of 3077 unique DON reports published from January 1996 to December 2023. The mean number of reports per year was 109.9 (SD 36.9). The highest number of reports occurred in 2014 with 204 DONs, of which 33% (68 out of 204) were reports on Influenza A and 30% (61 out of 204) were reports on Ebola. The lowest number of reports occurred in 2011, with 59 reports.

By month, the number of DON reports has fluctuated over time, with several noticeable peaks in the data. The highest peak in number of reports occurred in May 2009, for which 85% of reports (35 out of 41) were on the global Influenza A H1N1 pandemic. The second highest peak occurred in May 2003, for which 65% of reports (24 out of 37) were on the global severe acute respiratory syndrome coronavirus (SARS-CoV-1) outbreak. The third highest peak occurred in April 2014, for which 34% of reports (11 out of 32) were on the Ebola outbreak in Guinea, 31% of reports (10 out of 32) were on the Ebola outbreak in Liberia, and 31% of reports (10 out of 32) were on the Influenza A H7N9 outbreak in China. There were no reports published in December 1997 and January 2013. The mean number of reports published per month was 9.21 (SD 6.02). Fig 1 shows the overall trends by month for each transmission group.

**Fig 1.**
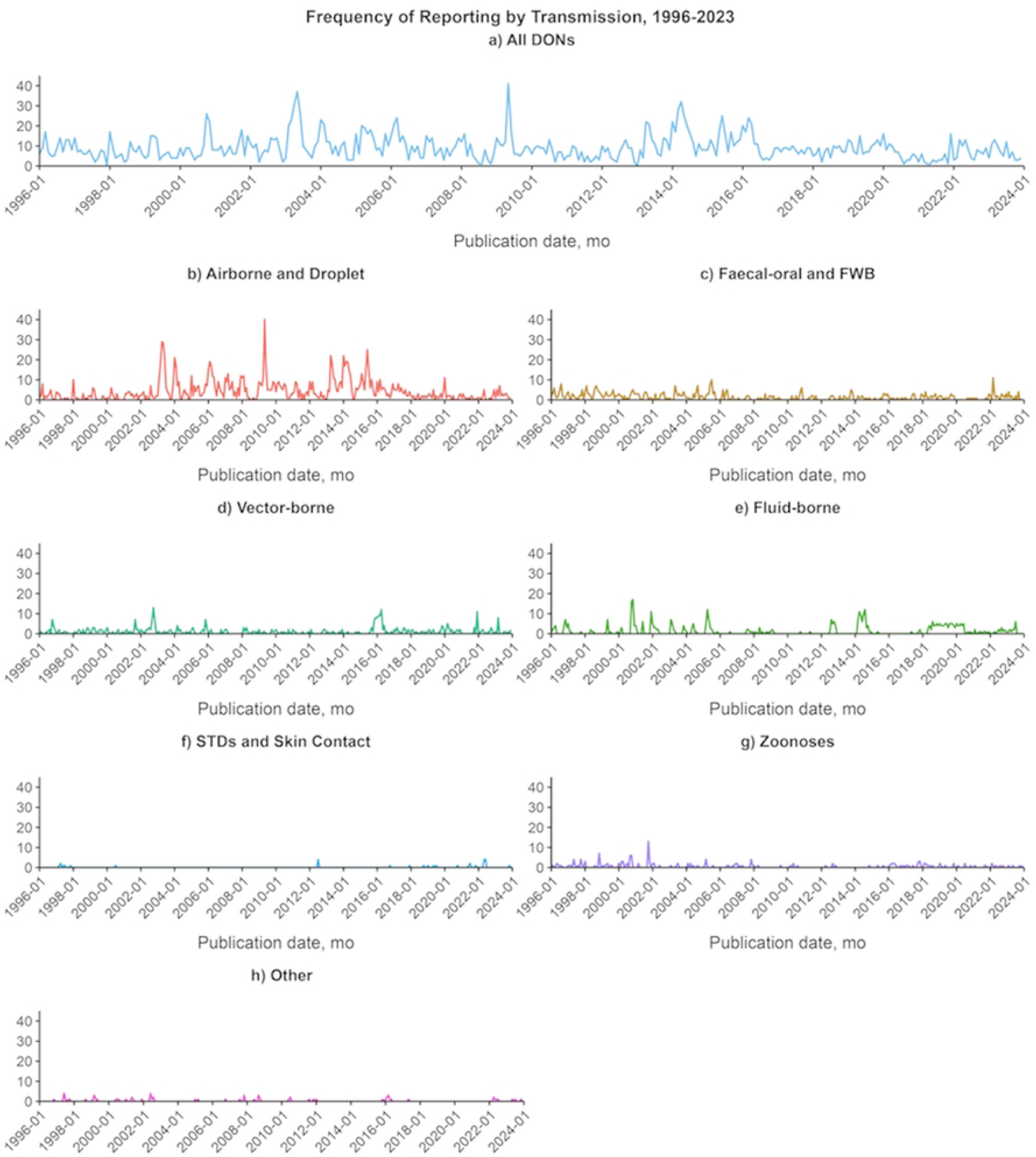
Frequency of reporting in the World Health Organization’s Disease Outbreak News (DONs), 1996-2023, by disease transmission group. No. reports are aggregated by month and year of publication. A) All DONs. B) Airborne and Droplet transmission-related DONs. C) Faecal-oral and FWB transmission-related DONs. D) Vector-borne transmission-related DONs. E) Fluid-borne transmission-related DONs. F) STDs and Skin Contact transmission-related DONs. G) Zoonoses. H) Other diseases. FWB, food-borne and water-borne diseases. STDs, sexually transmitted diseases.

Next, to understand seasonality in reporting, we aggregated DONs across 1996-2023 by calendar month and analyzed data for each transmission group (Fig 2). For all DONs and Airborne and Droplet transmission-related DONs, reporting in the latter half of the year (July-December) is lower than reporting in earlier months (January-June).

**Fig 2.**
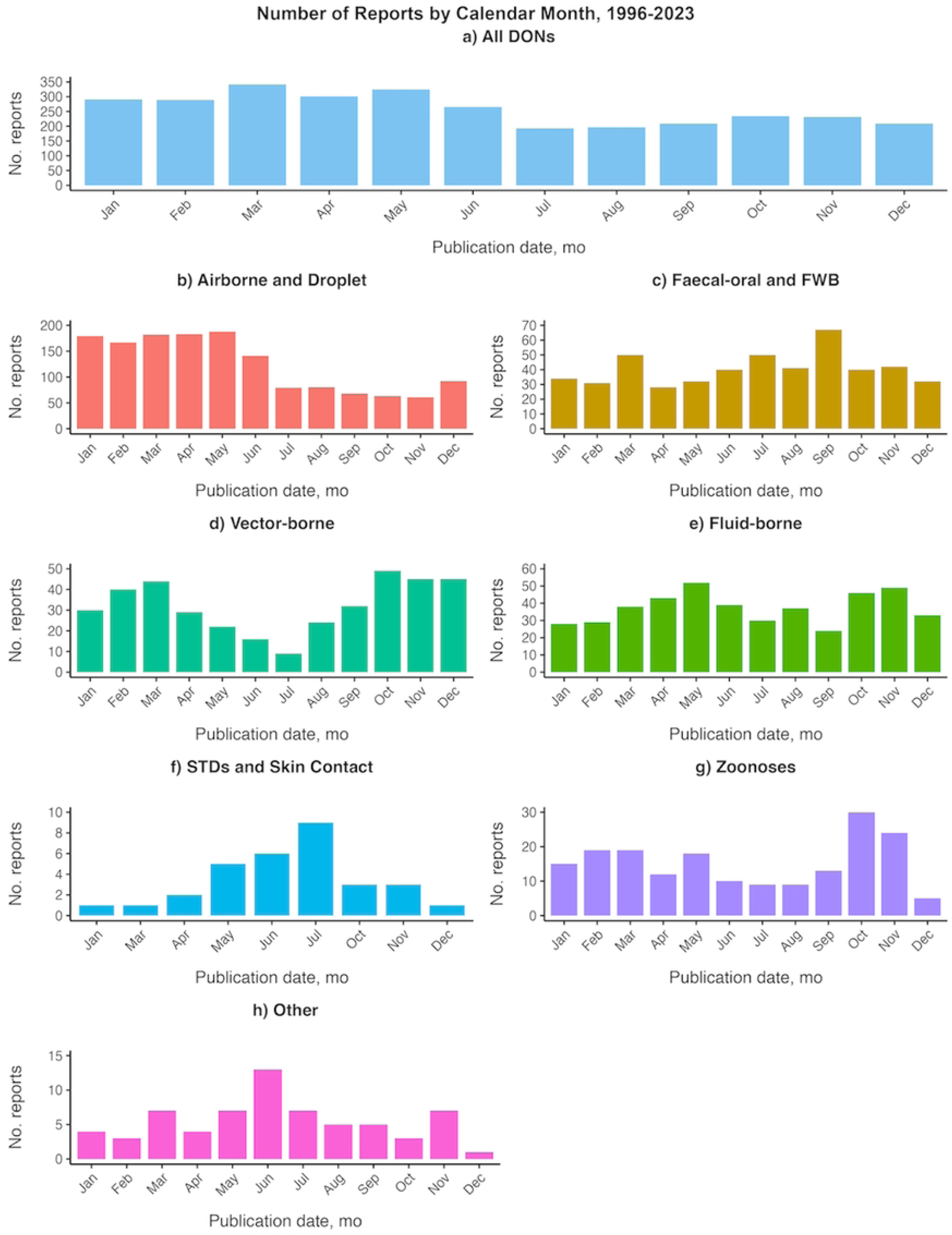
Frequency of reporting in the World Health Organization’s Disease Outbreak News (DONs), 1996-2023, by calendar month and disease transmission group. A) All DONs. B) Airborne and Droplet transmission-related DONs. C) Faecal-oral and FWB transmission-related DONs. D) Vector-borne transmission-related DONs. E) Fluid-borne transmission-related DONs. F) STDs and Skin Contact transmission-related DONs. G) Zoonoses. H) Other diseases. FWB, food-borne and water-borne diseases. STDs, sexually transmitted diseases. FWB, food-borne and water-borne diseases.

To understand how reporting varies between hemispheres, we aggregated DON reports across 1996-2023 and compared the number and percentage of reports by calendar month between the Northern and Southern hemispheres (Fig 3). For all DONs, in each calendar month, the proportion of reports distributed between the Northern and Southern hemispheres is similar, with the Northern hemisphere representing an average of 71.58% of reports, and the Southern hemisphere representing an average of 28.42% (Fig 3b).

**Fig 3.**
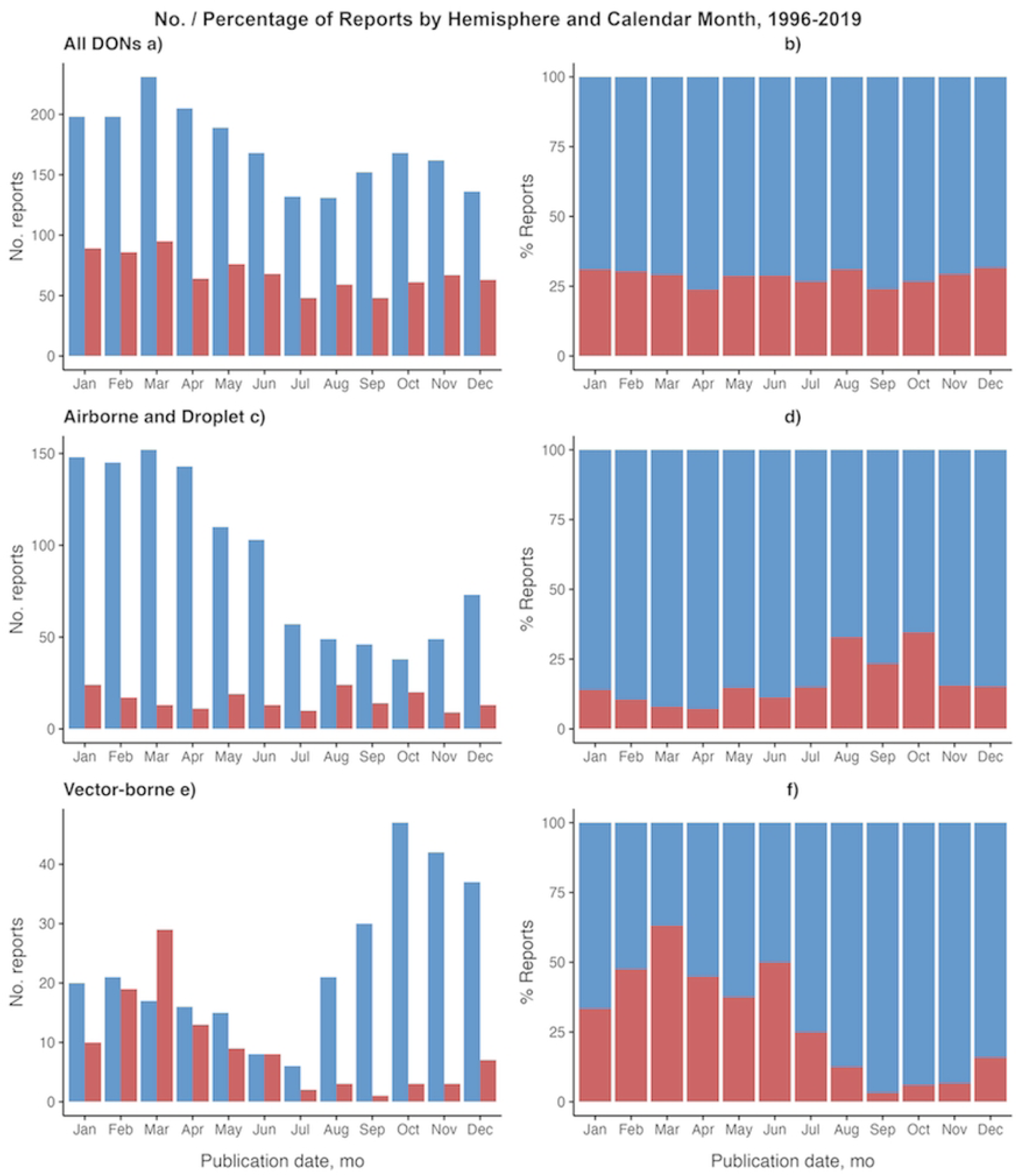
Reporting distribution of the World Health Organization’s Disease Outbreak News (DONs) reports, by Northern and Southern hemisphere. A) No. reports for all DONs. B) % reports for all DONs. C) No. reports for Airborne and Droplet transmission-related DONs. D) % reports for Airborne and Droplet transmission-related DONs. F) No. reports for Vector-borne transmission-related DONs. G) % reports for Vector-borne transmission-related DONs.

When analyzing specific transmission groups, we observed that the count and proportion of reports from each hemisphere vary across time. The Northern Hemisphere has significantly higher report counts than the Southern Hemisphere. Notable differences between hemispheres were observed for Airborne and Droplet transmission-related DONs and Vector-borne transmission-related DONs. For Airborne and Droplet transmission-related diseases, the Northern hemisphere peaked in the earlier month of March with 152 reports over 1996-2023, and saw the lowest number of 38 reports in October. On the other hand, the Southern hemisphere peaks in the later months of August and October, with 24 and 20 reports, which represent a proportion of 32.88% and 34.48% of total reports in the month, respectively. The Southern hemisphere saw the lowest levels of 11 reports (7.14%) in April.

For Vector-borne diseases, differences in reporting between hemispheres are more significant. The Northern hemisphere peaked in October, with 47 reports across 1996-2023, and fell to its lowest of 6 reports in July, while the Southern hemisphere reached its peak in March with 29 reports (63.04%), outnumbering the report count in the Northern hemisphere, and fell to its lowest of only 1 report (3.22%) in September.

Opposite patterns in seasonality are observed for Airborne and Droplet transmission-related DONs and Vector-borne transmission-related DONs, reflecting the contrasting seasons in each hemisphere (Fig 3). We excluded other transmission groups with very low levels of report counts in the Southern hemisphere from this analysis, as minor variations in reporting could skew the results.

### Time Series Analysis

#### By Disease Transmission Group

Time series analysis was performed on all DONs and the following disease transmission groups with sufficient data: Airborne and Droplet, Faecal-oral and FWB, Fluid-borne, and Vector-borne. Three disease transmission groups were not modelled due to low levels of data (STDs and Skin Contact, Zoonoses, Other). Negative binomial regression models were selected as the final models for all DONs and three disease transmission groups (Airborne and Droplet, Faecal-oral and FWB, and Vector-borne). No final model was selected for the Fluid-borne disease transmission group, as the negative binomial regression did not converge, and the Poisson and linear regression models did not yield statistically significant results.

Across all DONs, no significant trend in reporting was detected, and there was an annual pattern of seasonality (12 months). Similarly, for Airborne and Droplet transmission-related DONs, no significant trend in reporting was detected, and there was an annual pattern of seasonality (12 months). For DONs on diseases of Faecal-oral and FWB transmission, a significant negative trend in reporting was detected (p < 0.001, coefficient = −1.06e-04, 95% CI [−1.47e-04, −6.57e-05]), where the number of reports is expected to decrease by −0.13% every year (−0.0106% per month). In addition, 9-month seasonality in reporting was detected. For DONs on diseases of Vector-borne transmission, no significant trend was detected, and there was an annual pattern of seasonality (12 months). The final negative binomial regression models are shown in Fig 4. Tables of the negative binomial regression results are included in S8 Table.

**Fig 4.**
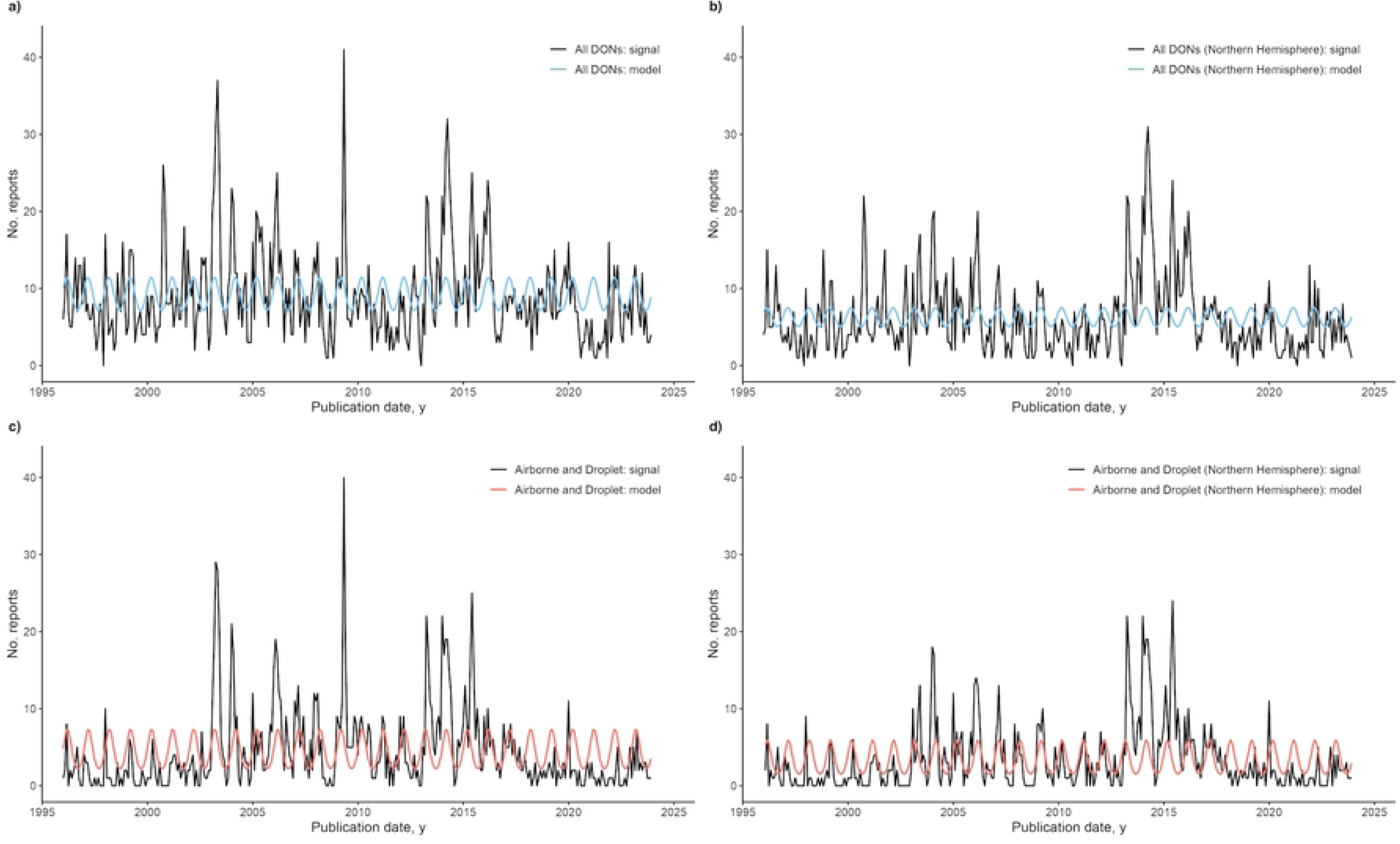
Negative binomial regression models of the World Health Organization’s Disease Outbreak News (DONs) reports. No. reports are aggregated by month and year of publication. Prior to model fitting, DONs for each disease transmission group were analyzed for trend and periodicity. A) Signal and negative binomial regression model for all DONs. For all DONs, no trend was observed. The model included seasonality of 12 months. B) Signal and negative binomial regression model for Airborne and Droplet transmission-related DONs. No trend was observed. The model included seasonality of 12 months. C) Signal and negative binomial regression model for Faecal-oral and FWB transmission-related DONs. Negative trend was observed, and the model included seasonality of 9 months. D) Signal and negative binomial regression model for Vector-borne transmission-related DONs. No trend was observed, and the model included seasonality of 12 months. FWB, food-borne and water-borne diseases.

### By Hemisphere

Next, we compared model results for all DONs and DON reports concerning the Northern hemisphere for each transmission group. As only the Airborne and Droplet transmission group had sufficient data after the hemisphere split, other transmission groups were excluded from the analysis. For all DONs, a similar model fit was observed for DONs in the Northern hemisphere as compared to total DONs before hemisphere split. For Airborne and Droplet transmission-related DONs of the Northern hemisphere, no significant trend was detected, and there was an annual pattern of seasonality (12 months). Comparing the models of Airborne and Droplet transmission-related DONs, a change in the coefficients of the sine and cosine terms resulted in an increase in the amplitude from 0.60 to 0.68. Thus, there was an increase in seasonality for Airborne and Droplet transmission-related DONs of the Northern hemisphere. The final model results are shown in Fig 5, and a table of the results is included in S9 Table.

**Fig 5.**
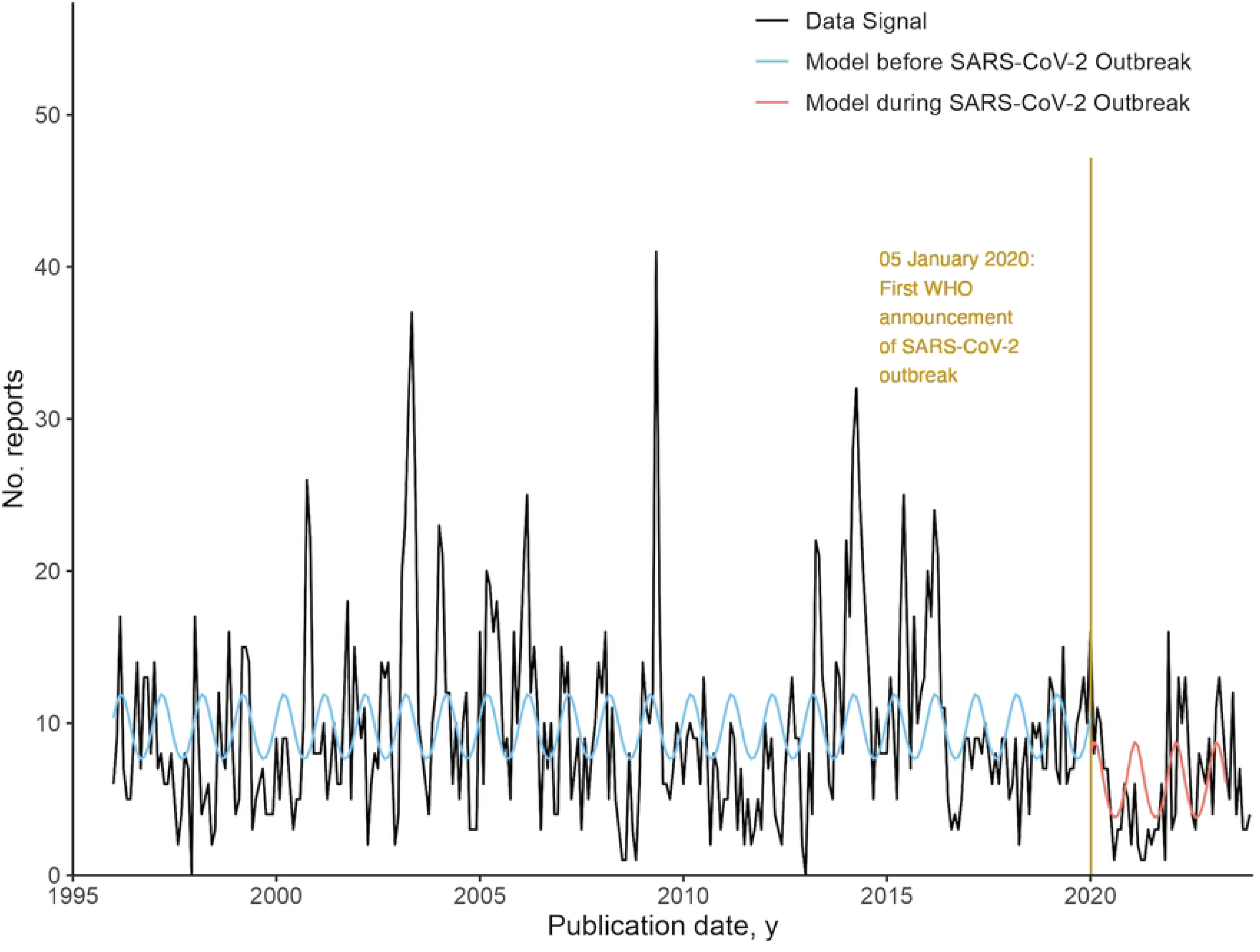
Negative binomial regression models of the World Health Organization’s Disease Outbreak News (DONs) reports, for all DONs and DONs of the Northern hemisphere only. Prior to model fitting, DONs for each group were analyzed for trend and periodicity. A) Signal and negative binomial regression model for all DONs. No trend was observed, and seasonality of 12 months was included. B) Signal and negative binomial regression for all DONs of countries in the Northern hemisphere. No trend was observed, and seasonality of 12 months was included. C) Signal and negative binomial regression model for Airborne and Droplet transmission-related DONs. No trend was observed, and seasonality of 12 months was included in the model. D) Signal and negative binomial regression model for Airborne and Droplet transmission-related DONs of countries in the Northern hemisphere. No trend was observed and seasonality of 12 months was included in the model.

### Interrupted time series analysis, before and during the COVID-19 pandemic

The final models selected for the interrupted time series analysis of reporting across all DONs before and during the COVID-19 pandemic were negative binomial regression models of report count against seasonality of 12 months. For both before and during COVID-19 pandemic models, no significant trend was detected. There is a step-change difference of - 3.73 reports at the baseline where predictor variables are equal to zero for both models. The results are shown in Fig 6, and a table of the results is included in the S10 Table.

**Fig 6.**
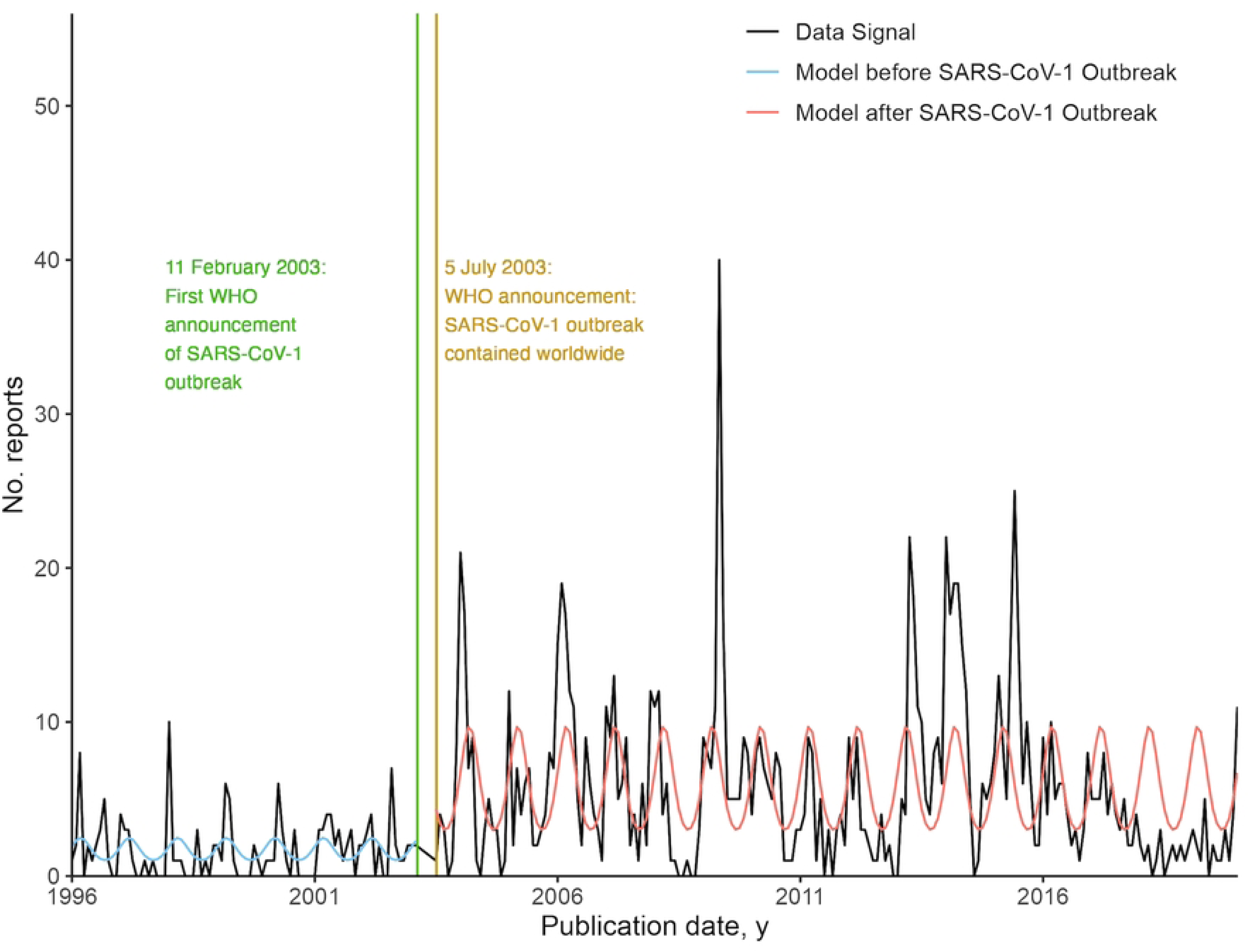
Negative binomial regression models of all Disease Outbreak News (DONs) reports, before and during the COVID-19 pandemic.

### Interrupted time series analysis, before and after the 2003 SARS-CoV-1 outbreak

From visual inspection of the data signal of Airborne and Droplet-related DONS, higher amplitudes and more frequent peaks were observed after the first significant peak of SARS-CoV-1 reports (27 reports) in May 2003. We investigated changes in reporting behavior after the SARS-CoV-1 outbreak, through interrupted time series analysis of Airborne and Droplet-related DONs, before and after the outbreak.

The final models selected for the interrupted time series analysis before and after the 2003 SARS-CoV-1 outbreak were negative binomial regression models of report count against seasonality of 12 months. For both before and after SARS-CoV-1 models, no significant trend was detected. There is a step-change difference of 3.77 reports at the baseline where predictor variables are equal to zero for both models. The results are shown in Fig 7, and a table of the results is included in S11 Table.

**Fig 7.**
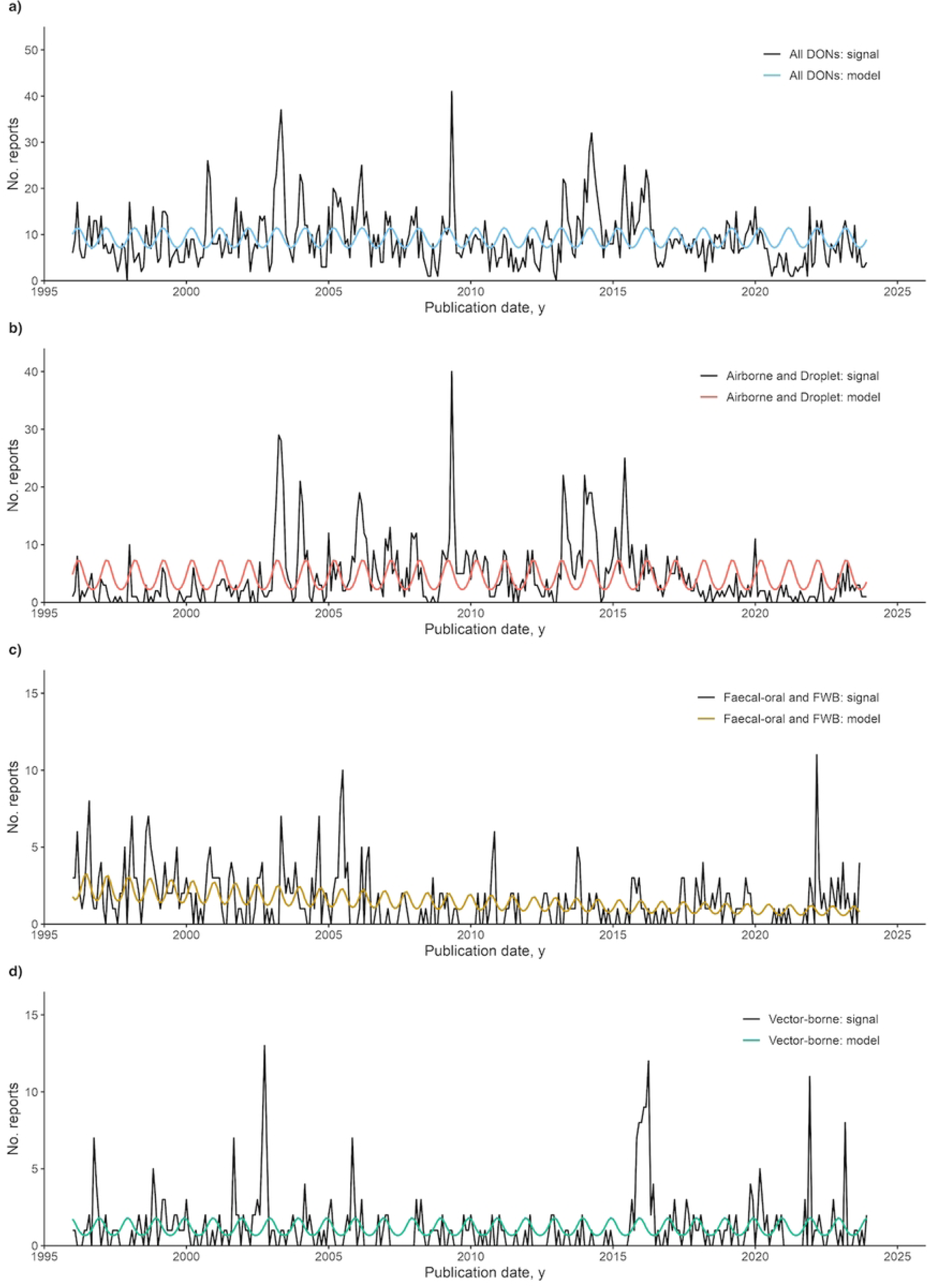
Negative binomial regression models of Airborne and Droplet transmission-related Disease Outbreak News (DONs) reports, before and after the 2003 SARS-CoV-1 outbreak.

## Discussion

Our analysis elucidated reporting patterns in the DON, across all DONs and by disease transmission group. For all DONs and Airborne and Droplet transmission-related DONs, reporting in the earlier half of the year (January-June) was higher than reporting in the latter half of the year (July-December). Comparing reporting in the Northern and Southern hemispheres, report count was higher for the Northern hemisphere. For all DONs and models for Airborne and Droplet and Vector-borne transmission groups, no significant trend was detected, and annual seasonality of 12 months was observed. Faecal-oral and FWB transmission-related DONS showed a significant negative trend and a seasonality pattern of 9 months.

In addition, we have observed that the occurrence of significant outbreaks, such as the COVID-19 pandemic and the 2003 SARS-CoV-1 outbreak, can affect reporting in the DONs. During the COVID-19 pandemic, reporting decreased across all DONs, with a step-change difference of −3.73 reports at baseline, comparing models before and during the pandemic.

This result supports the observations by Weets et al. (2025) [14], that the DONs were infrequently used to track the COVID-19 pandemic. After the 2003 SARS-CoV-1 outbreak, reporting of Airborne and Droplet transmission-related DONs increased, with a step-change difference of 3.77 reports at baseline, comparing models before and after the outbreak.

Our results evidence that reporting biases such as seasonality in reporting exist in the DONs. Understanding the reporting behavior of the DON can help the WHO, global health researchers and public health officials understand how the DON may be best utilized for outbreak management and research. There are several limitations in using the DONs and the DON database as a data source. The DONs and the DON dataset do not include every disease outbreak in history and should not be assumed to be a representative database of global disease outbreaks. As a collection of press releases, outbreaks published in the DON are determined by the WHO on a case-by-case basis. Not every outbreak will generate a DON, and outbreaks which require frequent updates, such as a public health emergency of international concern (PHEIC), may generate multiple DONs. Researchers should be aware of the low report counts observed across disease transmission groups such as ‘STDs and Skin Contact’, ‘Zoonoses’ and ‘Other’. Researchers should also note that variables on case numbers and deaths were not included this study due to missing data; preliminary inspection of the dataset of DONs across 1996-2019 revealed that 9% of all observations (313 observations) did not have information on case numbers, and 24% of all observations (817 observations) did not have information on deaths [13]. The dataset of DONs across 2020-2023 did not include variables on case numbers and deaths [14].

For future research on the DONs, it could be more beneficial to focus on each hemisphere or on specific regions to achieve more accurate results, as seasonal differences in reporting can complicate the model fitting and weaken seasonality. With the rich amount of textual data in each report, researchers can also consider alternative ways by which the analysis of the DONs can contribute to global health research. For instance, large language models (LLMs) could be used to analyze text data, particularly in historical or archival research to understand global health governance and shifts in global disease surveillance.

Beyond the DON dataset developed by Carlson et al. (2023) and current efforts of the WHO to maintain the DON as an authoritative source of information on disease outbreaks, the need for verified information and the potential of the DON to be used for retrospective and predictive analysis calls for the development of an indexable, global database of verified disease outbreaks. Developing a structured database that can be readily used for data analysis would complement current prose-based methods of outbreak news dissemination through the DON and the Weekly Epidemiological Record. While independent outbreak surveillance platforms such as HealthMap, ProMED-mail, GIDEON and global.health have demonstrated usefulness in the timely sharing of verified information, in the light of post-pandemic efforts to strengthen global collaboration against disease outbreaks, we propose that WHO consider the development of a structured global disease outbreak database, which would further streamline information dissemination and improve health emergency preparedness as an easily accessible, trusted resource for researchers and policymakers globally.

## Data Availability

All data are publicly available.

## Acknowledgments

This work was conducted as a dissertation project, in partial fulfillment of a Master of Philosophy (MPhil) degree at the University of Cambridge.

## Supporting Information

**S1 Table. List of R packages used.**

**S2 Table. Observations of DONid “DON-1996-01-22-a”, with relevant variables selected to illustrate that a single report can contain information on outbreaks in different countries.**

**S3 Table. Observations of DONid “DON-2010-10-25b”. In this Disease Outbreak News (DON) report, two disease outbreaks (Crimean-Congo haemorrhagic fever and dengue fever) were reported in Pakistan.**

**S4 Table. An example of observations in the original DON dataset with duplicate links and different DONids. The incorrect entry was marked with a “Y” in the “Exclude” column and removed from the final dataset.**

**S5 Table. Selected variables of interest which contain general information about the DON reports.**

**S6 Table. Selected examples of DONs showing the difference between DiseaseLevel1 and DiseaseLevel2.**

**S7 Table. The list of diseases featured in the DON dataset, by disease transmission group.**

**S8 Table. Final negative binomial regression results for all DONs and DONs of the following transmission groups: Airborne and Droplet, Faecal-oral and FWB, and Vector-borne diseases.**

**S9 Table. Final negative binomial regression results for all DONs and Airborne and Droplet transmission-related DONs, showing a comparison in model results for DONs before the hemisphere split and DONs of the Northern hemisphere.**

**S10 Table. Final negative binomial regression results for all DONs, before and during the COVID-19 pandemic.**

**S11 Table. Final negative binomial regression results for Airborne and Droplet transmission-related DONs, before and after the 2003 SARS-CoV-1 outbreak.**

